# Structural differences in adolescent brains can predict alcohol misuse

**DOI:** 10.1101/2022.01.31.22269833

**Authors:** Roshan Prakash Rane, Evert Ferdinand de Man, JiHoon Kim, Kai Görgen, Mira Tschorn, Michael A. Rapp, Tobias Banaschewski, Arun L.W. Bokde, Sylvane Desrivières, Herta Flor, Antoine Grigis, Hugh Garavan, Penny Gowland, Rüdiger Brühl, Jean-Luc Martinot, Marie-Laure Paillère Martinot, Eric Artiges, Frauke Nees, Dimitri Papadopoulos Orfanos, Herve Lemaitre, Tomáš Paus, Luise Poustka, Juliane H. Fröhner, Lauren Robinson, Michael N. Smolka, Jeanne Winterer, Robert Whelan, Gunter Schumann, Henrik Walter, Andreas Heinz, Kerstin Ritter, IMAGEN Consortium

## Abstract

Alcohol misuse during adolescence (AAM) has been linked with disruptive structural development of the brain and alcohol use disorder. Using machine learning (ML), we analyze the link between AAM phenotypes and adolescent brain structure (T1-weighted imaging and DTI) at ages 14, 19, and 22 in the IMAGEN dataset (*n* ∼ 1182). ML predicted AAM at age 22 from brain structure with a balanced accuracy of 78% on independent test data. Therefore, structural differences in adolescent brains could significantly predict AAM. Using brain structure at age 14 and 19, ML predicted AAM at age 22 with a balanced accuracy of 73% and 75%, respectively. These results showed that structural differences preceded alcohol misuse behavior in the dataset. The most informative features were located in the white matter tracts of the corpus callosum and internal capsule, brain stem, and ventricular CSF. In the cortex, they were spread across the occipital, frontal, and temporal lobes and in the cingulate cortex. Our study also demonstrates how the choice of the phenotype for AAM, the ML method, and the confound correction technique are all crucial decisions in an exploratory ML study analyzing psychiatric disorders with weak effect sizes such as AAM.

## 1. INTRODUCTION

Many adolescents participate in risky and excessive alcohol consumption behaviors [1], especially in European and North American countries. Several studies have identified that such early and risky exposure to alcohol is a potential risk factor that can lead to the development of Alcohol Use Disorder (AUD) later in life [2, 3, 4]. During adolescence and early adulthood (age 10-24), the human brain undergoes maturation characterized by an increase in white matter (WM) [5] and an initial thickening and later thinning of grey matter (GM) regions [6]. Researchers have suggested that excessive alcohol use during this period might disrupt normal brain maturation, causing lifelong effects [1, 7, 8]. Therefore, understanding how alcohol misuse during adolescence is related to the development of Alcohol Use Disorder (AUD) later in life is crucial to understanding alcohol addiction. Furthermore, uncovering how adolescent alcohol misuse (AAM) is associated with the adolescent brain at different stages of its development can help to implement a more informed public health policy surrounding alcohol use during this age.

### Previous studies

Several studies in the last two decades have attempted to uncover how adolescent alcohol misuse (AAM) and their structural brain are related. These are summarised in Table S1 in the supplementary text. Most of the earlier studies collected data from small but controlled groups of 30 to 100 subjects and compared specific brain regions such as the hippocampus or the pre-frontal cortex (pFC) between adolescent alcohol misusers (AAMs) and mild users or non-users (controls). They used structural features such as regional volume [9, 10, 11], cortical thickness [12], or white matter tract volumes [13, 14]. These studies found differences between the groups in regions such as the hippocampus [9, 10], cerebellum [11], and the frontal cortex [11]. However, these findings are not always consistent across studies [15]. This is also evident from the highlighted texts in our literature review in Table S1. Another group of studies attempted to uncover if alcohol misuse disrupts the natural developmental trajectory of adolescent brains [16, 17, 18, 14, 19, 20]. As compared to controls, these studies reported that the brains of AAMs showed accelerated GM decline [17, 18, 19] and attenuated WM growth [17, 19]. However, brain regions reported were not consistent between these studies either and do not tell a coherent story [15] (see Table S1). These differences in findings could potentially be due to the following reasons:

1. **Heterogeneous disease with a weak effect size:** Alcohol misuse has a heterogeneous expression in the brain [21]. This heterogeneity might be driven by alcohol misuse affecting diverse brain regions in different sub-populations depending on demographic, environmental, or genetic differences [22]. Furthermore, the effect of alcohol misuse on adolescent brain structure can be weak and hard to detect (especially with the mass-univariate methods used in previous studies). The possibility of several disease sub-types exasperated by the small signal-to-noise ratio can generate incoherent findings regarding which brain regions are affected by alcohol.
2. **Higher risk of false-positives:** Most previous studies have small sample size that are prone to generate inflated effect size [23]. Furthermore, these studies employ mass-univariate analysis techniques that are vulnerable to *multiple comparisons problem* [24] and can produce false-positives if ignored. These factors coupled with the possibility of publication bias to produce positive results [25] can have a high likelihood of generating false-positive findings [26].
3. **Several metrics to measure alcohol misuse:** There is no consensus on what is the best phenotype to measure AAM. Many studies use binge drinking or heavy episodic drinking as a measure of AAM [12, 27, 14, 20], while few others use a combination of binge drinking, frequency of alcohol use, amount of alcohol consumed and the age of onset of alcohol misuse [28, 18, 29, 30, 19]. These differences in the analysis could potentially produce different findings.

### Multivariate exploratory analysis

Over the last years, data collection drives such as IMAGEN [31], NCANDA [32], and UK Biobank [33] made available large-sample multi-site data with *n >* 1000 that are representative of the actual population. This enabled researchers to use multivariate, data-driven, and exploratory analysis tools such as machine learning (ML) to detect effects of alcohol misuse on multiple brain regions [27, 34, 30]. Such whole-brain multivariate methods are preferable over the previous mass-univariate methods as they have a higher sensitivity to detect true positives [35]. Furthermore, ML can be easily used for clinical applications such as computer-aided diagnosis, predicting future development of AUD, and future relapse of patients into AUD [36].

Due to these advantages, several exploratory studies using ML have been attempted in AUD research [27, 30, 34]. We further extend this line of work by analyzing the newly available longitudinal data from IMAGEN (*n* ∼ 1182 at 4 time points of adolescence) [31] by designing a robust and reliable ML pipeline. The goal of this study is to explore the relationship between adolescent brain and AAM using ML on a relatively large (*n* ≥ 1000), multi-site adolescent data and discover the brain regions associated with AAM. As shown in Figure 1, we predict AAM at age 22 using brain morphometrics derived from structural imaging captured from three stages of adolescence – ages 14, 19, and 22. The structural features of different brain regions are extracted from two modalities of structural MRI, that is, T1-weighted imaging (T1w) and Diffusion Tensor Imaging (DTI). The most informative structural features for the ML model prediction are visualized using SHAP [37, 38] that reveals the most distinct structural brain differences between AAMs and controls. Furthermore, we use multiple phenotypes of alcohol misuse such as the frequency of alcohol consumption, amount of consumption, onset of misuse, binge drinking, the AUDIT score, and other combinations, and systematically compare them. We also compare four different ML models, and multiple methods of controlling for confounds in ML and derive important methodological insights which are beneficial for reliably applying ML to psychiatric disorders such as AUD. To promote reproducibility and open science, the entire codebase used in this study, including the initial data analysis performed on the IMAGEN dataset are made available at *https://github.com/RoshanRane/imagenml*.

**Figure 1.**
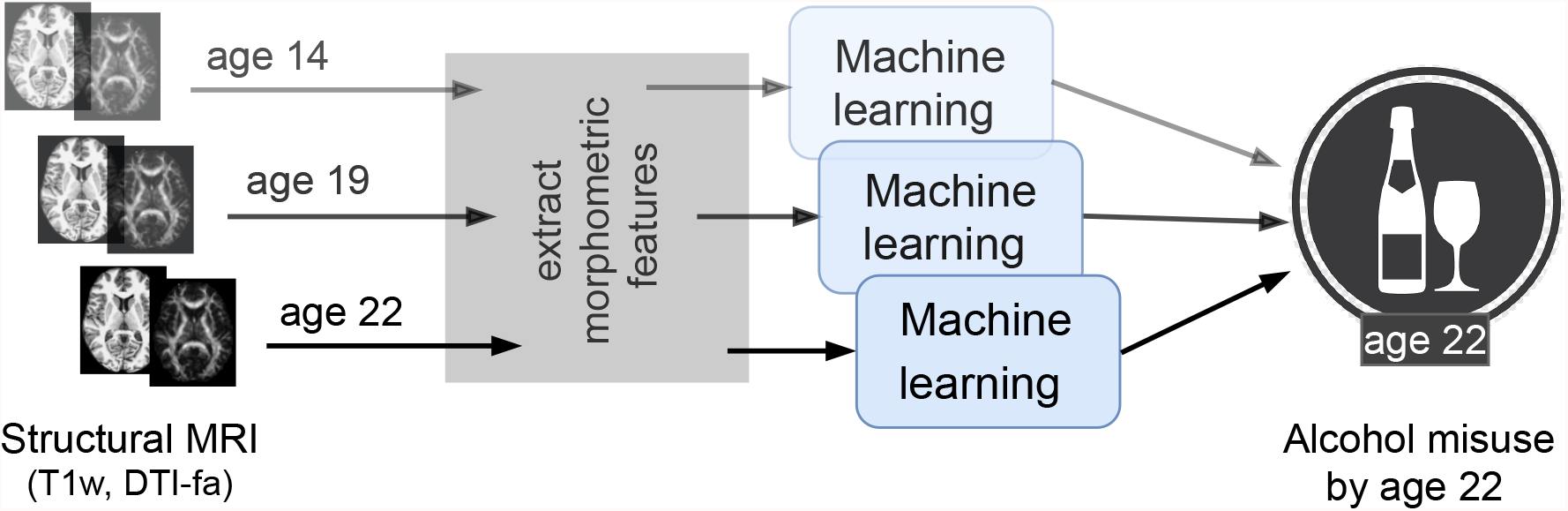
An overview of the analysis performed. Morphometric features extracted from structural brain imaging are used to predict Adolescent Alcohol Misuse (AAM) developed by the age of 22 using machine learning. To understand the causal relationship between AAM and the brain, three separate analyses are performed by using imaging data collected at three stages of adolescence: age 14, age 19 and age 22

## 2 DATA

The IMAGEN dataset [31, 39] is currently one of the best candidates for studying the effects of alcohol misuse on the adolescent brain. Most large-sample studies listed in Table S1 [27, 29, 30] used the IMAGEN dataset for their analysis. It consists of data collected from over 2000 young people and includes information such as brain neuroimaging, genomics, cognitive and behavioral assessments, and self-report questionnaires related to alcohol use and other drug use. The data was collected from 8 recruitment centers across Europe, at 4 successive time points of adolescence and youth. Figure 2 (a) shows the number of subjects at each time point and the number of participants that were scanned. Subjects were not scanned in FU1. More details regarding recruitment of subjects, acquisition of psychosocial measures, and ethics can be found on the IMAGEN project website^1^.

**Figure 2.**
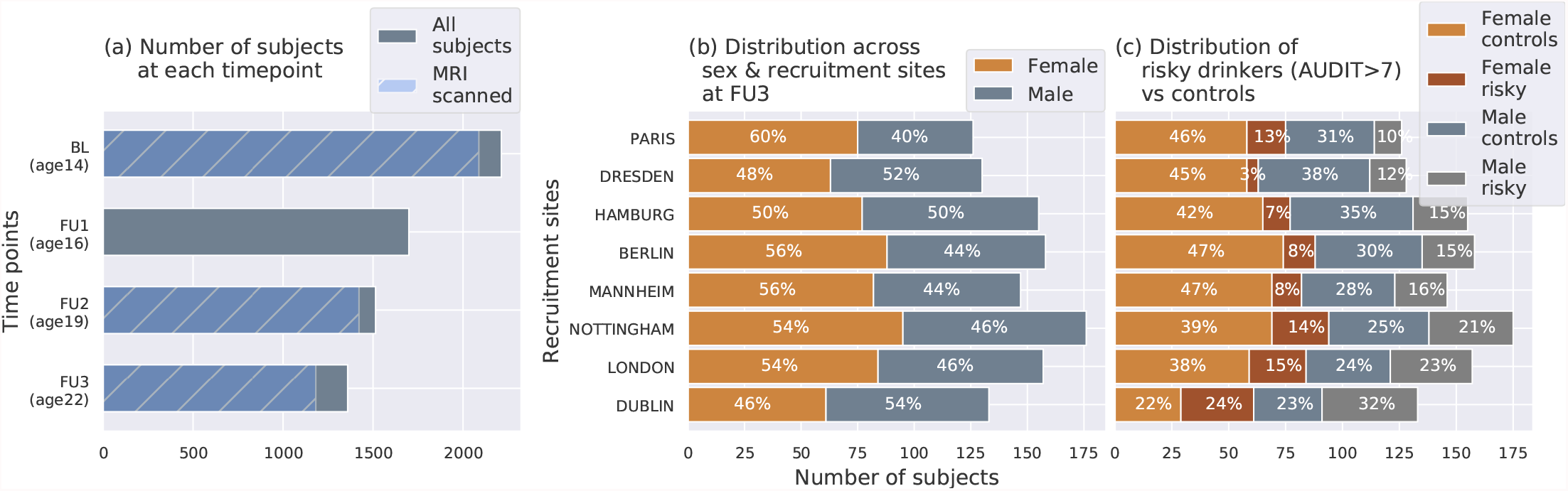
The IMAGEN dataset: (a) Data is collected longitudinally at 4 stages of adolescence - age 14 or baseline (BL), age 16 or follow-up 1 (FU1), age 19 or follow-up 2 (FU2) and, finally age 22 or follow-up 3 (FU3). The blue bar shows the number of subjects with brain imaging data. (b) The distribution of subjects across sex and the site of recruitment, for the 1182 subjects that were scanned at FU3 (c) The same distribution across sex and site also showing the proportion of subjects that meet the AUDIT ‘risky drinkers’ category at FU3.

### Structural neuroimaging data

To investigate the effects of alcohol on brain structure, two MRI modalities have been used predominantly in the literature - (a) T1-weighted imaging (T1w), and (b) Diffusion Tensor Imaging (DTI) (see Table S1). While T1w MRI can be used to derive general features of the brain structure such as cortical and sub-cortical volumes, areas, and gray-matter thicknesses, DTI captures white matter microstructures by probing water molecule motion. An axial slice (*z* = 80) of both of these MRI modalities of a control subject from the IMAGEN data are shown in Figure 3.

**Figure 3.**
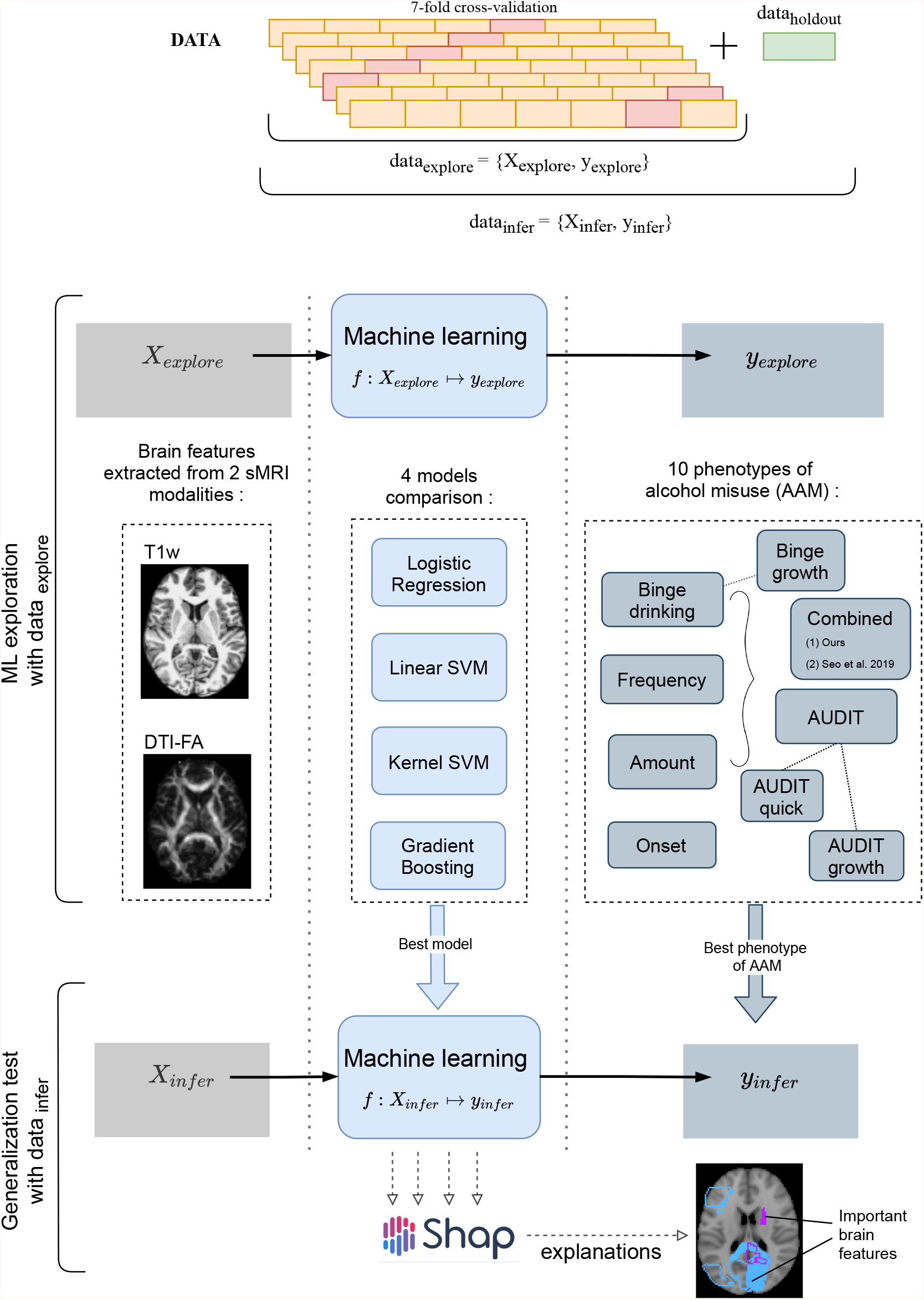
A schematic representation of the experimental procedure. followed for all 3 time point analyses. In the ML exploration stage, we experiment with four ML models and 10 phenotypes of AAM on 80% of the data (data_*explore*_) using a 7-fold cross validation scheme. Once the best ML model, the best phenotype of AAM, and the most appropriate confound-control technique are determined, the *generalization test* is performed on data_*infer*_ by using the data_*holdout*_ subset as the test data. The result from the generalization test are reported as the final results and the informative brain features are determined at this stage using SHAP [37].

Both modalities were recorded using 3-Tesla scanners. The T1w images were collected using sequences based on the ADNI protocol [40]. The IMAGEN consortium used Freesurfer’s recon-all pipeline to process these images and extract structural features. This involves registering the T1w-images to the Talairach template brain, automatic extraction of gray matter, white matter and cerebrospinal fluid (CSF) sections, and then segmenting them into 34 cortical regions per hemisphere and 45 sub-cortical regions.The grey matter volume (in mm^3^), surface area (in mm^2^), thickness (in mm), and surface curvature, are extracted for each of the cortical regions using the Desikan-Killiany atlas, along with global features such as total intracranial, total grey matter, white matter and CSF volumes. For the subcortical regions, the mean intensity and volume are determined. This results in a total of 656 structural features per subject. DTI scans were acquired using the protocol described in Jones et al. [41] and Fractional Anisotropy (FA) is derived from the DTI using FMRIB’s Diffusion Toolbox FDT. The DTI-FA images are then non-linearly registered to the MNI152 space (1mm^3^) and the average FA intensity at 63 regions with white matter tracts are calculated using the TBSS toolbox [42] by the IMAGEN consortium^2^. Subjects with FA intensity greater than 3 standard deviations from the mean are excluded as outliers.

### Alcohol misuse phenotypes

Information related to alcohol use and misuse can be found in the AUDIT screening test^3^ (Alcohol Use Disorder Identification Test), ESPAD questionnaire (European School Survey Project on Alcohol and other Drug), and the TLFB logs (Timeline-Followback Interview). Previous studies used different metrics of alcohol misuse such as the number of binge drinking episodes [12, 27, 14, 20], the frequency and amount of alcohol consumption [28, 18, 29, 30, 19], and even the age of onset of alcohol misuse [43] to characterize AAM. There has not yet been a systematic comparison of these different phenotypes.

In this paper, we use four alcohol misuse metrics to derive ten phenotypes of AAM, (a) frequency of alcohol use, (b) amount of alcohol consumed per drinking occasion, (c) year of onset of alcohol misuse, and (d) the number of binge drinking episodes. These phenotypes are listed in Table 1 and include each of the individual metrics, their combinations, and their longitudinal trajectories from age 14 to 22. The longitudinal phenotypes, ‘Binge-growth’ and ‘AUDIT-growth’, are generated using latent growth curve models [44] to capture the alcohol misuse trajectory over the four time points - BL, FU1, FU2, and FU3. To derive the AAMs group and the controls from each alcohol misuse metric, a standard procedure is followed that is similar to Seo et al. [30] and Ruan et al. [43]. First, the phenotype is used to categorize the subjects into three stages of alcohol misuse severity - heavy AAMs, moderate misusers, and safe users. Moderate misusers are then excluded from the analysis (250 400 subjects) and ML classification is performed with heavy misusers as AAMs and safe users as controls. Figure S2 and Table S2 in the supplement shows how the subjects are divided into these three sub-groups for each of the 10 phenotype and also lists the final number of subjects in each sub-group in the FU3 analysis, as an example. The data analysis procedure can be found in the project code repository^4^ within the *dataset-statistics* notebook.

**Table 1.** 10 phenotypes of Adolescent Alcohol Misuse (AAM) are derived and compared in this analysis. A description of each phenotype is provided here along with the link to the IMAGEN questionnaires ID used to generate the phenotype.

| No. | Phenotype | Description | Questionnaire |
| --- | --- | --- | --- |
| 1 | Frequency | Number of occasions drinking alcohol in last 12 months | ESPAD 8b. |
| 2 | Amount | Number of alcohol drinks consumed on a typical drinking occasion | ESPAD prev31, AUDIT q2. |
| 3 | Onset | Had one or more binge-drinking experiences by the age of 14 | ESPAD 29d |
| 4 | Binge | Total drunk episodes from binge-drinking in lifetime (by age 22) | ESPAD 19a, AUDIT q3. |
| 5 | Binge-growth | Longitudinal trajectory of binge-drinking experiences had per year | Growth curve of ESPAD 19b. |
| 6 | AUDIT | AUDIT screening test performed at the year of scan | AUDIT-total (q1-10). |
| 7 | AUDIT-quick | Only the first 3 questions of AUDIT screening test | AUDIT-freq (q1-3). |
| 8 | AUDIT-growth | Longitudinal changes in the AUDIT score measured over the years | Growth curve of AUDIT-total. |
| 9 | Combined-seo | A combined risky-drinking phenotype from Seo et al. [30] generated using amount, frequency, and binge-drinking data | ESPAD 8b, 17b, 19b, and TLFB alcohol2 |
| 10 | Combined-ours | A combined risky-drinking phenotype developed by clustering amount, frequency, and binge-drinking trajectory | AUDIT q1, q2, ESPAD 19a, growth curve of ESPAD 19b. |

### Confounds in the dataset

Diagram (c) in Figure 2 shows how the proportion of risky alcohol users varies across the 8 recruitment sites and among the male and female subsets at each site within the dataset. For example, a greater portion of subjects from sites like Dublin, London, and Nottingham indulge in risky alcohol use compared to the sites from mainland Europe. Similarly, at most sites, a greater portion of males are risky alcohol users compared to females. These systematic differences can confound ML analyses since ML models can use the sex and site information present in the neuroimaging data to indirectly predict AAM, instead of identifying alcohol-related effects in the brain structure. This problem of confounds in multivariate analysis [45, 46, 47] and the methods used to control for its effects are explained in further detail in the next section.

## 3 METHODS

Three time point analyses are performed in this study. Each time point analysis is divided into two stages called the *ML exploration* stage and the *generalization test* stage. The ML exploration is performed with 80% of data (randomly sampled). The remaining 20% (*n* = 147) serve as an independent test data, called the data_*holdout*_, which is only used once, in the end, to perform the final inference and report the results. This design allows us to first determine the best ML algorithm for the task and the best phenotype of AAM, and then test the results on an independent subset of the data. The pseudocode of this procedure is also provided in the supplementary section 4. It was implemented with the help of python’s *scikit-learn* software package^5^. The two-stage cross validation (CV) with a inner n-fold cross validation (CV) procedure is designed to prevent ‘double dipping’ [48, 49]. All data preprocessing and analysis is executed only on the training data in data_*explore*_, and only applied on the test data during validation. This ensures that there are no data leakage issues that were found in several previous ML neuroimaging studies [50].

### MRI features

The 656 morphometric features extracted from T1w sMRI modality and the 63 features extracted from the DTI-FA modality are used together as the input for the ML models at both stages. Each feature is standardized to have zero mean and unit variance across all subjects (mean and variance are estimated only on the training data, and then applied to the test data). Features with zero variance are dropped.

### ML models

Four ML models are tested in this study. These include logistic regression (LR), linear SVM (SVM-lin) [51], kernel SVM with a radial basis function (KSVM-rbf) [52], and a gradient boosting (GB) classifier [53]. LR and SVM-lin are linear ML methods, whereas SVM-rbf and GB are capable of learning non-linear mappings. We use the liblinear [54] implementation of SVM-lin and XGBoost [55] implementation of GB. GB is an ensemble learning method. The hyperparameters of the models are listed in the supplementary section 3 are tuned using an inner-CV. Testing 4 different ML models helps to account for any modeling-related bias [56] in the final results. Combining the 4 ML models and the ten different phenotypes of AAM, we end up with a total of 40 ML classification runs in the ML exploration stage.

### Evaluation metrics

The model performance is evaluated using the *balanced accuracy* metric [57]. It is formulated as the mean of the model’s accuracies for each class (AAM and controls) in the classification. Therefore, it is insensitive to class imbalances in the data. Along with this, the area under the curve of the receiver-operator characteristic (AUC-ROC) is also calculated. In ML exploratory stage, 7 measures are obtained for each metric from the outer 7-fold CV which helps to estimate mean of the model performance and get a sense of the variance [58]. During generalization test, the ML models are retrained 7 times on data_*explore*_ with different random seeds and reevaluated on data_*holdout*_ to gain an estimate of the model performance on data_*holdout*_. The statistical significance of the final generalization test accuracies is calculated using permutation testing [59]. The permutation test is performed by running the entire ML pipeline with randomly shuffled labels in the training data, while keeping the labels in the test data fixed. This is repeated 1000 times to generate the null-hypothesis (*H*_0_) distribution and derive the p-value. Since three time point analyses are performed on the same subjects, Bonferroni correction is applied on the p-values to control for the false-positive rate from this multiple comparison.

### Model interpretation

The associations learned by the ML models between structural brain features and AAM is extracted using a post-hoc feature importance attribution technique called SHAP [37]. SHAP (SHapley Additive exPlanations) uses the concept of *Shapley Values* from cooperative game theory to fairly determine the marginal contribution of each input feature to model prediction [37].

Following the generalization test, a SHAP value (*S*_*s,f*_) is generated for each input feature *f* of each subject *s* in data_*holdout*_. The goal is to determine which of the 719 features were most informative for the model when classifying AAMs from controls. Feature importance can be determined by looking at the average absolute SHAP value of each feature across all subjects 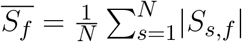, where *N* denotes the total subjects in data_*holdout*_. The most significant features are chosen as those features that have 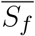 value at least two times higher than the average SHAP value across all the features 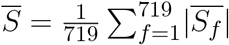. Since the generalization test is repeated seven times with different random seeds, we have seven instances of 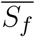 available. Only those features that consistently have 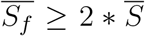 across all seven runs are listed as the most informative features. Next, it is determined if these informative features have higher-than-average or lower-than-average values when predicted as AAM. This information is further relevant for deriving clinical insights about how AAM brain structure differs from controls.

### Correcting for confounds

In ML, a confounding variable *c* is defined as a variable that correlate with the target *y* and is deducible from the input *X*, and this relationship *X* → *c* →*y* is not of primary interest to the research question and hinders the analysis [47]. As demonstrated by the diagram on the right, a confounding variable *c* can form an alternative explanation for the relationship between *X* and *y* and distract the ML models from detecting the signal of interest *s*_*y*_ between *X* → *y*. In this study, the sex of the subjects and their site of recruitment can confound the AAM analysis [30] since they correlate with the output AAM labels and are predictable from the input structural brain features. Instead of detecting the effects of alcohol misuse in the brain *s*_*y*_, the ML models could potentially use the information about the confounds *s*_*c*_ to predict AAM along the alternative pathway (shown with the red dotted lines) and produce significant but confounding results [30, 47, 60]. In neuroimaging studies, two methods have been extensively employed for correcting the influence of confounds:

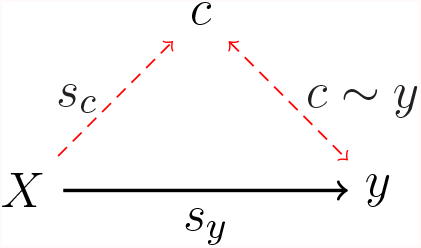

4 *Confound regression*: In this method, the influence of the confounding signal *s*_*c*_ on *X* is controlled by regressing out the signal from features in *X* [45]. This can remove the alternative confounding explanation pathway by eliminating the link *s*_*c*_ between *X* → *c*.
5 *Post hoc counterbalancing*: The correlation between the confound and the output *c* ∼ *y* can be removed by resampling the data after the data collection. This method potentially removes the alternative confounding pathway by abolishing the relationship *c* → *y* [45]. The resampling is performed such that the distribution of the values of the confounding variable *c* is similar across all classes of *y* (AAM and controls). So for example, after counterbalancing for sex in this study, the ratio of male-to-female subjects should be the same in AAMs and controls. One common technique of counterbalancing for categorical confounds (eg. sex, site) involves randomly dropping some samples from the larger classes in *y* until they are equal. This is called counterbalancing *with undersampling*. However, this will result in a reduction in the sample size and hence the statistical power of the study. Another way to counterbalance without losing samples involves performing *sampling-with-replacement* on the smaller classes in *y*. This is called counterbalancing *with oversampling*. One should take care that the sampling-with-replacement is done only on the training data, after the train-test split is performed.

To assess whether confound regression worked and the confounding signal *s*_*c*_ is removed successfully, a confound correction method recently proposed by Snoek et al. [47] can be used. In this method, the ML algorithm used in the original analysis is reused to predict the confound *c* from the neuroimaging data *X*. Following a successful confound regression, the confound should not be predictable anymore from *X* and *X* → *c* should produce insignificant or chance accuracy. Similarly, to determine if counterbalancing was successful and the correlation *c* ∼ *y* was removed, we used the *Same Analysis Approach* by Görgen et al. [46]. Here, the same ML algorithm is used to predict the confound *c* from the labels *y* [46]. An above-chance significant prediction accuracy between *c* → *y* would indicate that the correlation *c* → *y* still exists and the counterbalancing was not successful. Since the confounds *c*_*sex*_ and *c*_*site*_ are categorical, they are first one-hot encoded to ensure no false ordinal relationship is implied. The confound correction methods are only performed on the training data as recommended by Snoek et al. [47]. The balanced accuracy metric used ensures that we account for any class imbalances in the test data. Before starting the ML exploration, we first compare these different confound correction methods and choose the most suitable method among them.

## 4 RESULTS

The results are reported in the following four subsections: In subsection 1, different confound-control techniques are compared and the most suitable technique for this study is determined. Subsection 2 shows the results of the ML exploration performed with ten AAM labels, four ML models, and using imaging data from three time points of adolescence. This stage helps to determine the best phenotype of AAM and the best ML model. Subsection 3 reports the final results on the independent data_*holdout*_ for all three time point analyses and subsection 4 shows the most informative features found in each of the analyses.

### Confound correction techniques

The sex *c*_*sex*_ and recruitment site *c*_*site*_ of subjects confound this study (see section 2) and their influence on the study needs to be controlled. We test three confound correction techniques on data_*explore*_ – (a) confound regression (b) counterbalancing with undersampling and (c) counterbalancing with oversampling. To verify if these methods work as expected, the *same analysis approach* from Görgen et al. [46] and the approach by Snoek et al. [47] are employed. For the two confounds *c*_*sex*_ and *c*_*site*_, this requires us to test five input-output combinations (*X* → *y, X* → *c*_*sex*_, *X* → *c*_*site*_, *c*_*sex*_ → *y* and *c*_*site*_ → *y*) for a given *X* → *y* analysis.

Figure 4 shows the results of comparing different confound correction techniques for the ‘Binge’ phenotype. The following conclusions can be derived from this comparison:

**Figure 4.**
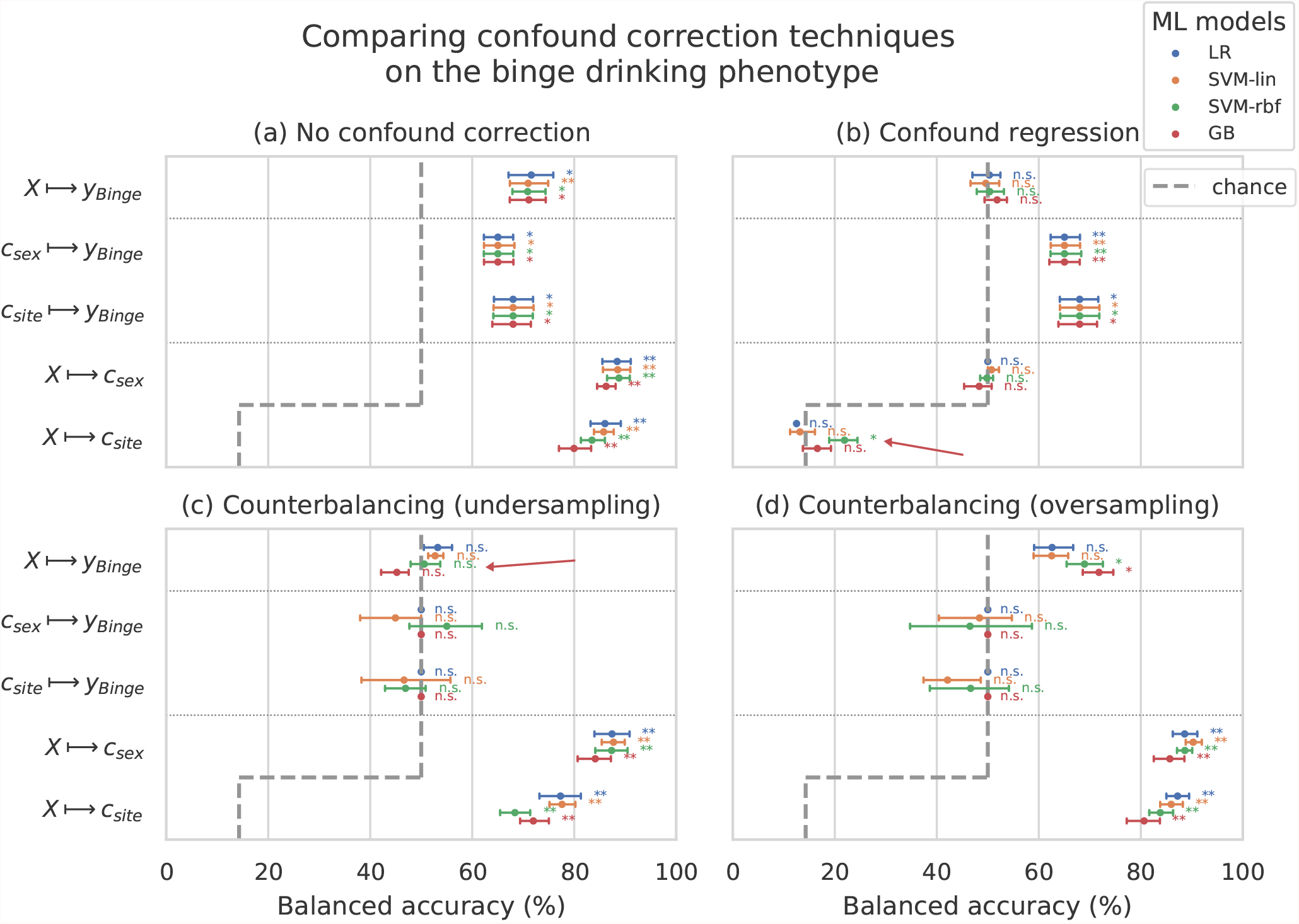
Comparing confound correction techniques. Five input-output settings are compared within each confound correction technique: *X* → *y, X* → *c*_*sex*_, *X* → *c*_*site*_, *c*_*sex*_ → *y*, and *c*_*site*_ → *y*. (a) shows the results before any correction is performed, (b) shows the results of performing confound regression, and (c) and (d) show the results from counterbalancing by undersampling the majority class and oversampling the minority class, respectively. Statistical significance is obtained from 1000 permutation tests and is shown with ** if *p <* 0.01, * if *p <* 0.05, and ‘n.s’ if *p* ≥ 0.05.

1. Sex and site can confound the AAM analysis: As shown in subplot (a), all the input-output combinations involving the confounds (*X* → *c*_*sex*_, *X* → *c*_*site*_, *c*_*sex*_ → *y* and *c*_*site*_ → *y*) produce significant prediction accuracies before any confound correction is performed. This further adds to the evidence that both the confounds *c*_*sex*_, *c*_*site*_ can strongly influence the accuracy of the main analysis *X* → *y* and confound the analysis.
2. Confound regression is not a good choice when followed by a non-linear ML method: Following confound regression, the results of *X* → *c*_*sex*_ and *X* → *c*_*site*_ should become non-significant as the signal *s*_*c*_ has been removed from *X*. However, it is seen that in some cases the non-linear models SVM-rbf and GB are capable of detecting the confounding signal *s*_*c*_ from the imaging data. The red arrow in the subplot (b) points out one such case in the example shown. This is not surprising as the standard confound regression removes linear components of the signal *s*_*c*_ but does not remove any non-linear components that might still be present in *X* [46, 60]. Furthermore, confound regression carries an additional risk of also regressing-out the useful signal in *X* that does not confound the analysis *X* → *y* but is a co-variate of both *c* and *y* [60].
3. Counterbalancing with oversampling is the best choice for this study: As expected, counterbalancing forces the *c*_*sex*_ → *y* and *c*_*site*_ → *y* accuracies to chance-level by removing the correlation between *c* ∼ *y* (subplots (c) and (d)). It can be seen that after the undersampled counterbalancing the results of the main analysis *X* → *y* also become non-significant as indicated by the red arrow in (c). This drastic reduction in performance is likely due to the reduction in the sample size of the training data by *n* 100 250 from undersampling. Therefore, counterbalancing with oversampling of the minority group is a better alternative compared to undersampling.

This comparison was also repeated for two other AAM phenotypes - ‘Combined-seo’ and ‘Binge-growth’ and the above findings were found to be consistent across all of them. Hence, counterbalancing with oversampling is used as the confound-control technique in the main analysis. When performing over-sampled counterbalancing, it is ensured that the oversampling is done only for the training data.

### ML exploration

The results from the ML exploration experiments are summarised in Figure 5. For the different AAM phenotypes, the balanced accuracies range between 45 ™ 73%. It must be noted that the results across different phenotypes are not directly comparable as each AAM phenotype classification task has a different sample size varying between ≈620 − 780. These differences in the number of samples (see Table S2 in the supplement) in the two classes AAM and controls could add additional variance in the accuracy. Nevertheless, some useful observations can be made from the consistenties across the three time point analyses in subplots (a), (b) and (c):

**Figure 5.**
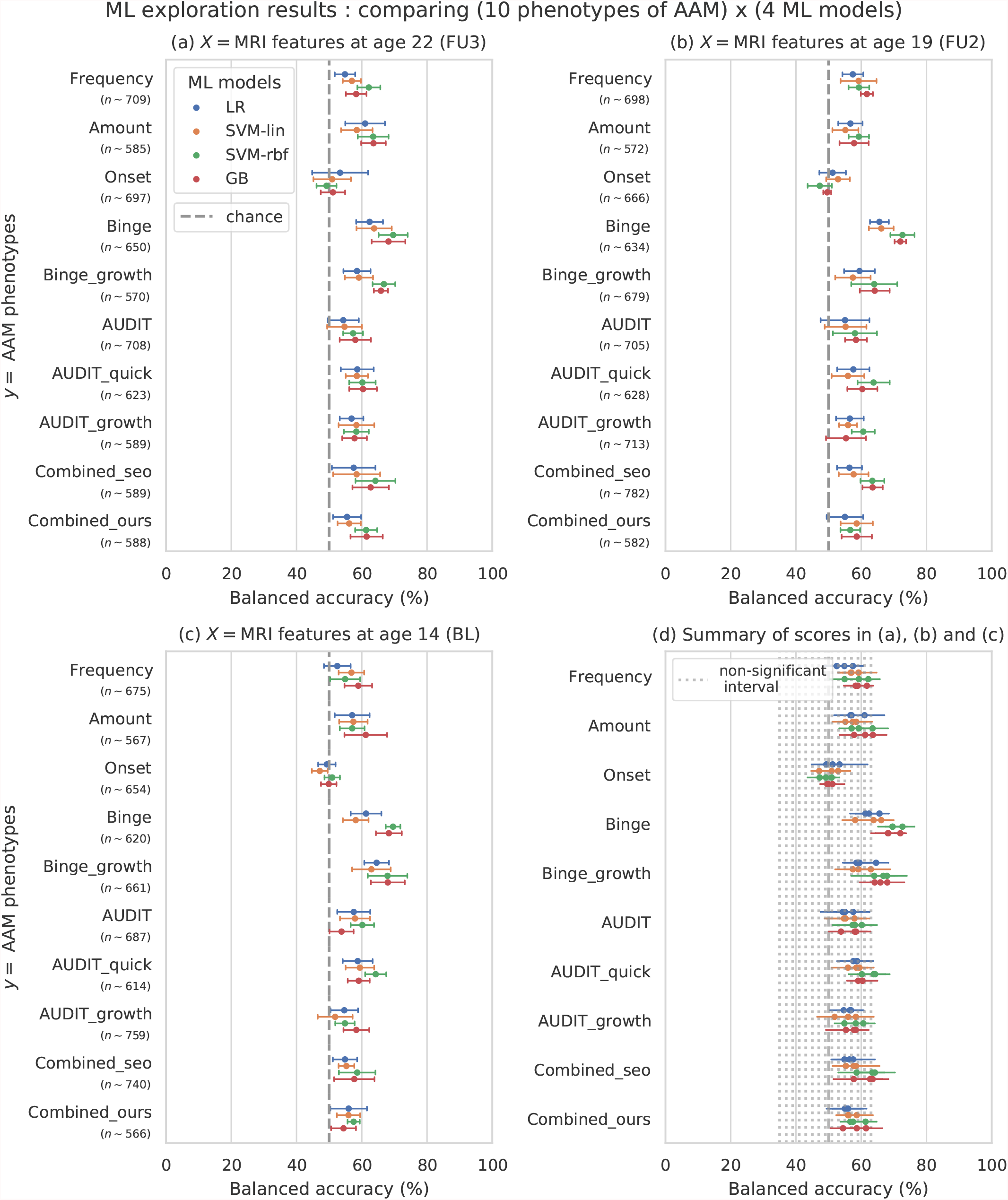
Results of the ML exploration experiments: The ten phenotypes of AAM tested are listed on the y-axis and the four ML models are represented with different color coding as shown in the legend of figure (a). For a given AAM label and ML model, the point represents the mean balanced accuracy across the 7-fold CV and the bars represent its standard deviation. Figure (a) shows the results when the imaging data from age 22 (FU3) is used, figure (b) shows results for age 19 (FU2) and figure (c) for age 14. Figure (d) shows the results from all three time point analyses in a single plot along with the interval of the balanced accuracy that were non-significant (*p* ≤ 0.05) when tested with permutation tests.

1. The most predictable phenotype from structural brain features for all three time point analyses is ‘Binge’ which measures the total lifetime experiences of being drunk from binge drinking.
2. Other individual phenotypes such as the amount of alcohol consumption (Amount), frequency of alcohol use (Frequency) and the age of AAM onset (Onset) are harder to predict from brain features compared to the binge drinking phenotype. The results on ‘Combined-seo’ and ‘Combined-ours’ shows that using these phenotypes in combination with binge drinking seems to also be detrimental to model performance.
3. All models perform poorly at predicting AAM phenotypes derived from AUDIT. This is surprising as AUDIT is considered a *de-facto* screening test for measuring alcohol misuse [61].
4. Among the four ML models, the SVM with non-linear kernel SVM-rbf, and the ensemble learning method GB perform better than the linear models LR and SVM-lin. This is further evident in the summary plot (d) in the figure.

In summary, the non-linear ML models SVM-rbf and GB coupled with the ‘Binge’ phenotype consistently perform the best in all three time point analyses. This is more clearly visible in the summary figure (d) where the results from all three analyses are combined in a single plot. Similar general observations can be made when the AUC-ROC metric is used to measure model performance as shown in the supplementary Figure S3.

### Generalization

The generalization test is performed with ‘Binge’ phenotype as the label and the two non-linear ML models, SVM-rbf and GB. The final results are shown in Figure 6. For the three analyses using imaging data from age 22, age 19, and age 14, as input, an average balanced accuracy of 78%, 75.5%, and 73.1% are achieved, respectively. Their average ROC-AUC scores are 83.9%, 83.1%, and 81.5% for the respective analyses. The accuracies for all three time point analyses are significant with *p <* 0.01. To get a better intuition, refer to the supplementary Figure S1 that shows model accuracies versus the accuracies obtained from permutation tests.

**Figure 6.**
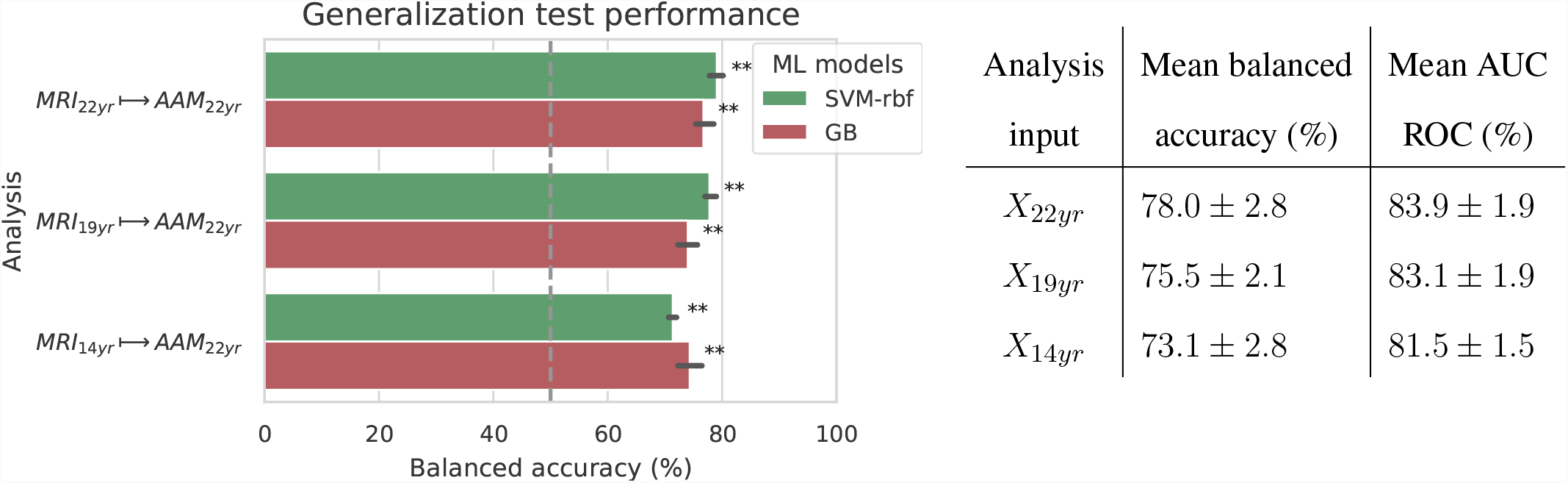
& Table 2. Final results for the three time point analyses on the ‘Binge’ drinking AAM phenotype obtained with the two non-linear ML models, kernel-based support vector machine (SVM-rbf) and gradient boosting (GB). The figure shows the mean balanced accuracy achieved by each ML model within each analysis while the table lists the combined average scores for each analysis. The ML models are retrained 7 times on data_*explore*_ with different random seeds and evaluated on data_*holdout*_ to obtain an estimate of the accuracy with a standard deviation. Statistical significance is obtained from 1000 permutation tests and is shown with ** if *p <* 0.01, * if *p <* 0.05, and ‘n.s’ if *p* ≥ 0.05.

### Important brain regions

Following the generalization test, the most informative structural brain features are determined for the SVM-rbf model, as it performs relatively better among the two non-linear models tested on data_*holdout*_ (see Figure 6). Figure 7 lists the most important features for all three time point analyses and shows whether these features have lower-than-average or higher-than-average values whenever the ML model predicts alcohol misuse.

**Figure 7.**
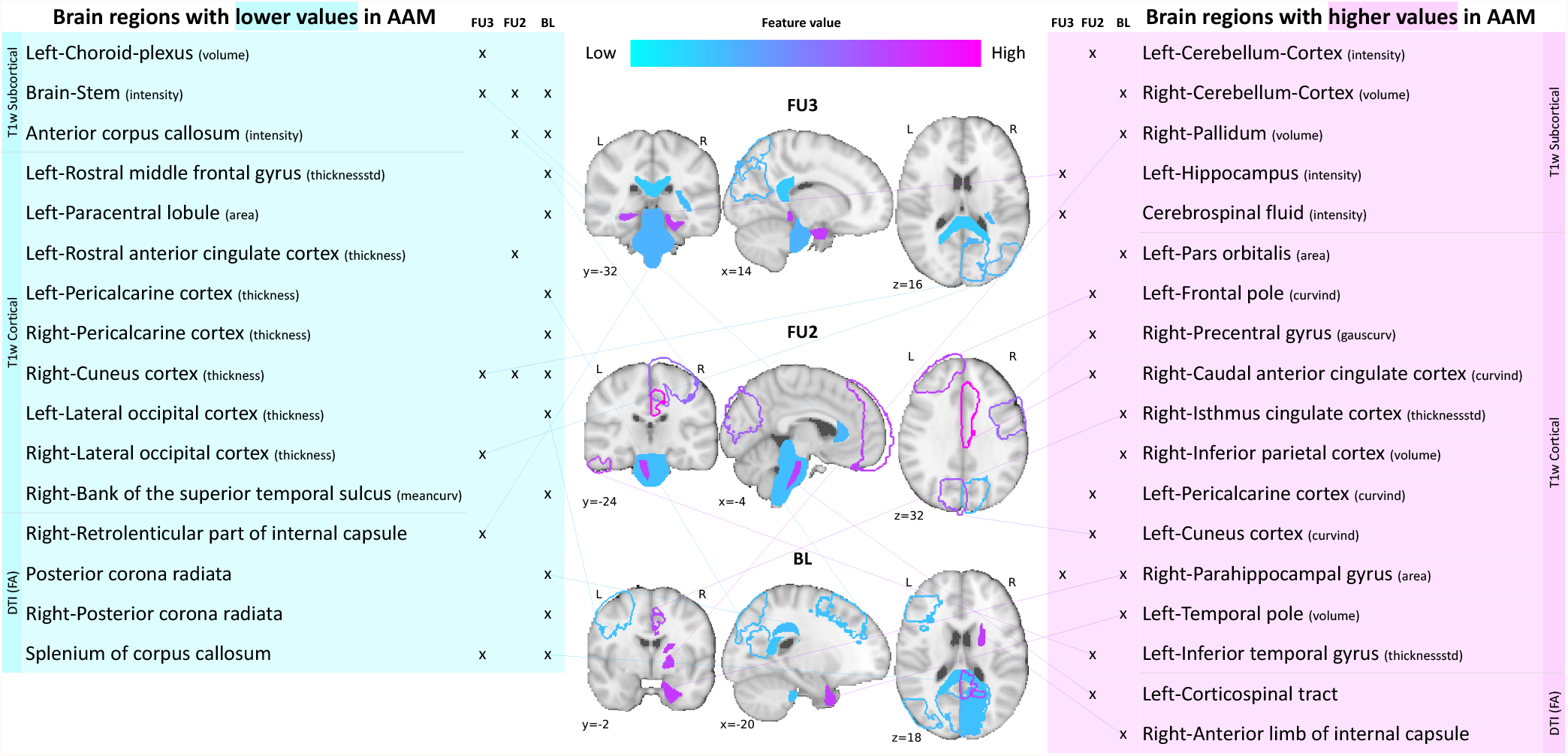
Most informative structural features for SVM-rbf model’s predictions on data_*holdout*_. All important features are listed and their locations are shown on a template brain for a better intuition for each of the three time point analyses. The features are color coded to also display whether these features have lower-than-average or higher-than-average values when the model predicts alcohol misusers. (Acronyms:: AAM: adolescence alcohol misuse, area: surface area, volume: gray matter volume, thickness: average thickness, thicknessstd: standard deviation of thickness, intensity: mean intensity, meancurv: integrated rectified mean curvature, gauscurv: integrated rectified gaussian curvature, curvind: intrinsic curvature index)

Several clusters of regions and feature values can be identified. Most of the important subcortical features are located around the lateral ventricles and the third ventricle and include CSF-related features such as the CSF mean-intensity, volume of left choroid plexus, and left corticospinal tract in the brain stem. Several white matter tracts are found to be informative such as parts of the corpus callosum, internal capsule, and posterior corona radiata. Furthermore, all of these white matter tracts, along with the brain stem have lower-than-average intensities in AAM predictions. The prominent cortical features are spread across the occipital, temporal, and frontal lobes. In the *MRI*_*age*22_ → *AAM*_*age*22_ analysis important cortical features appear in the occipital lobe. In contrast, for the future prediction analyses *MRI*_*age*19_ → *AAM*_*age*22_ and *MRI*_*age*14_ → *AAM*_*age*22_, clusters appear in the limbic system (parts of the cingulate cortex and right parahippocampal gyrus), frontal lobe (left-pars orbitalis, left-frontal pole, right-precentral gyrus, and left-rostral middle frontal gyrus) as well as in the temporal lobe (left-inferior temporal gyrus, left-temporal pole, and right-bank of the superior temporal sulcus). In the occipital lobe, AAMs predictions have lower grey matter thickness in the right-cuneus, lateral occipital, and pericalcarine cortices, and higher curvature index in left-cuneus and left-pericalcarine cortex.

## 5 DISCUSSION

For over two decades, researchers have tried to uncover the relationship that exist between adolescent alcohol misuse (AAM) and brain development. Many previous studies found that such a relationship exists (see Table S1) but with low-to-medium effect size [10, 27, 34, 30, 11, 13, 17]. The brain regions linked with AAM varied greatly across studies (see highlighted text in Table S1). This inconsistency in findings and effect sizes could be due to methodological limitations, small sample studies, unavailability of long-term longitudinal data like IMAGEN [31], or simply due to the heterogeneous expression of AAM in the brain. In our study, ML models predicted AAM with significantly above-chance accuracies in the range 73.1% − 78% (ROC-AUC in 81.5% − 83.9%) from adolescent brain structure captured at ages 14, 19 and 22. Thus, our results demonstrate that adolescent brain structure is indeed associated with alcohol misuse during this period.

The causality of the relationship between adolescent brain structure and AAM is not clear [27, 20]. The relationship could arise from alcohol misuse inducing neurotoxicity [21] causing the observed changes in their brains. It could also be that these structural differences precede AAM and such adolescents are just more vulnerable towards alcohol misuse [8, 62]. Such neuropsychological predisposition could stem from genetic predispositions or from influencing environmental factors such as early stress or childhood trauma [63, 64], misuse of other drugs such as cannabis [65] and tobacco, and parental drug misuse [14]. There might also be an interaction effect between alcohol-induced neurotoxity and environmental and genetic predispositions [20]. While the direction of causality is still under active investigation [20, 66], the significantly high accuracies obtained in our study for *MRI*_*age*19_ → *AAM*_*age*22_ and especially *MRI*_*age*14_ → *AAM*_*age*22_ suggest that these structural differences might be preceding alcohol misuse behavior. Out of the 265 subjects that took the ESPAD survey at age 14 and belonged to the AAM category in *MRI*_*age*14_ → *AAM*_*age*22_ analysis, 83.3% of subjects reported having no or just one binge drinking experience until age 14. This supports the results from Robert et al. [20] that a cerebral predisposition - be it due to genetic or environmental effects - might be preceding alcohol abuse in adolescents.

We identified the most informative brain features for the ML predictions using SHAP that has been successfully applied to medical data [37, 38]. The important features were found to be distributed across several subcortical and cortical regions of the brain, implying that the association between AAM and brain structure is widespread and heterogeneous. In accordance with previous studies, AAM was associated with lower DTI-FA intensities in several white matter tracts and the brain stem [13, 16, 15] and reduced GM thickness [34, 18], especially in the occipital lobe. Features of anterior cingulate cortex [34, 30, 15], middle frontal and precentral gyrus [17], hippocampus [9, 10], and right parahippocampal gyrus [27] were also found to be informative, although the type of feature and the average feature value in AAMs differed from previous studies. Features from the frontal lobe and cerebellum were informative only for future AAM [14] but not for current AAM prediction, in contrast to findings of [11, 27, 30]. This difference could be due to the meticulous confound control performed in this study for sex and site of the subjects. Additionally, our ML models also found CSF-related features in the third and lateral ventricles, and some regions of the temporal cortex as informative features for AAM prediction.

In the ML exploration stage, we found that the binge drinking phenotype, which is commonly used in previous studies [10, 27, 20], was the most predictable phenotype of AAM as compared to frequency, amount, or onset of alcohol misuse. Curiously, phenotypes derived from AUDIT, which is a gold standard of screening for alcohol misuse [61], did not score significantly above-chance in any of the three time point analyses. Other similar compound metrics that use measures of alcohol use frequency and amount along with binge drinking, such as ‘Combined-seo’ and ‘Combined-ours’, also perform worse than using just the binge drinking information. This suggests that using other phenotypes of alcohol misuse in combination with binge drinking was detrimental to the prediction task, as compared to using only binge drinking. Different phenotypes of AAM capture slightly different psychosocial characteristics of adolescents [67]. For instance, ‘Amount’ correlates significantly with agreeableness and a life history of relocation valence (*r* = −0.14, *p <* 0.001), accident valence (*r* = −0.16, *p <* 0.001) and sexuality frequency (*r* = −0.17, *p <* 0.001), whereas the other phenotypes do not (*p >* 0.01). ‘AUDIT’ and it’s derivatives significantly correlate with impulsivity trait (*r* = 0.23, *p <* 0.001) on SURPS, where as ‘Binge’ does not (*r* = 0.09, *p >* 0.01) but they both correlate with sensation seeking trait (*r >* 0.29, *p <* 0.001) as also found in previous studies [68]. Castellanos-Ryan et al. [68] have found that these two traits manifest differently in the brain. Therefore, one can hypothesize that the psychosocial differences (and their associated neural correlates [68]) between ‘Binge’ and the other AAM phenotypes might explain the 2 − 10% higher accuracy obtained with ‘Binge’.

### Methodological insights

To the best of our knowledge, this is the first study to analyze and reports results on the complete longitudinal data from IMAGEN, including the follow-up 3 data. Two previous studies, Whelan et al. [27] and Seo et al. [30] performed similar ML analysis on the IMAGEN data and unlike us, found only a weak association between structural imaging and AAM. The logistic regression model in Whelan et al. [27] scored 58 ± 8% ROC-AUC when predicting AAM at age 14 from structural imaging features collected at age 14 (BL) and 63 ± 7% ROC-AUC at predicting AAM at age 16 (FU1). This lower accuracy with high variance obtained in their experiments can be attributed to - (a) the relatively smaller sample size used in their study (*n* ∼ 265 − 271), (b) unavailability of long-term AAM information from IMAGEN’s FU2 and FU3 data, (c) using only a linear ML model, and (d) only using GM volume and thickness as structural features. On the other hand, Seo et al. [30]‘s models achieved accuracies in the range 56 − 58% when predicting AAM at age 19 (FU2) using imaging features from age 19, and did not get a significant accuracy when they used imaging features from age 14. This lower performance can be attributed to the following experimental design decisions - (a) Seo et al. [30] used GM volume and thickness features from just 24 regions of the brain associated to cue-reactivity, (b) their AAM phenotype is not the best phenotype of AAM as evident from the results of our ML exploration (see results for ‘Combined-seo’ in Figure 5), and (c) the confound-control technique used in their study, confound regression, can result in under-performance as demonstrated in Figure 4.

In contrast to these previous works, our study has the following advantages: First, we use 719 structural features extracted from 2 MRI modalities, T1w and DTI, that include not only GM volume and thickness but also surface area, curvature, and WM and GM intensities from all cortical and sub-cortical regions in the brains. Second, we empirically derive the best AAM label for the task by comparing different phenotypes previously used in the literature. For the different AAM phenotypes, the balanced accuracies range between chance to significant performance (45% − 73%), emphasizing the importance of the choice of the label in such ML studies with low effect sizes. And finally, we test different confound correction techniques and use the one that effectively controls for the influence of confounds without also destroying the signal of interest. In summary, the higher accuracy in the current study can be attributed to not just the availability of long-term data on AAM but also to the rigorous comparison of different labels of AAM, different ML models and confound control techniques.

Among the four different ML models tested, the two non-linear models, SVM-rbf and GB, consistently performed better than the two linear models. We also explicitly ensured that the confounding influence of sex and site were eliminated by combining suggestions from Görgen et al. [46] and Snoek et al. [47]. We found evidence that the linear confound regression technique used often in previous ML-based neuroimaging studies [30, 20, 47], might not be the best choice as it cannot be used with non-linear models such as SVM-rbf or Naive Bayes used in Seo et al. [30] and distorts the signal of interest from the neuroimaging data [60] as seen in Figure 4. In contrast, counterbalancing using oversampling is recommended as it successfully removed the influence of the confounds without reducing the sample size in the study.

In contrast to the main results, the models failed to achieve significant prediction in the *leave-one-site-out* experiment and the scores displayed high variance (refer supplement Figure S4). This variance could be caused by the widely distributed sample sizes across each site resulting in uneven folds in the n-fold CV (Figure 2). The chance performance might also be due to any site-specific variations in the data_*holdout*_ that prevailed despite the rigorous data acquisition standards enforced across the sites by the IMAGEN group [39].

### Future work

An important future work would be to understand how AAM correlates with several psycho-socio-economic variables and uncover any environmental risk factors such as childhood abuse, parental drug use, and life event stressors that could be mediating the relationship we discovered between AAM and brain structure. It would also be interesting to investigate if the functional connectivity (fMRI) in adolescent brains can also predict AAM [43]. Another important future work would be to reproduce the results on another data set comprising adolescents from a different geographic area such as NCANDA [32].

## 6 CONCLUSION

This study analyzed alcohol misuse in adolescents and their brain structure in the large, longitudinal IMAGEN dataset consisting of *n* ∼ 1182 healthy adolescents [39, 31]. We found that alcohol misuse in adolescents can be predicted from their brain structure with a significant and high accuracy of 73% 78%. More importantly, alcohol misuse at age 22 could be predicted from the brains at age 14 and age 19 with significant accuracies of 73.1% and 75.5%, respectively. This suggests that the structural differences in the brain might at least partly be preceding alcohol misuse behavior [20]. In contrast to previous large-sample studies that use ML [27, 30], we extensively compared different phenotypes of alcohol misuse such as frequency of alcohol use, amount of use, the onset of alcohol misuse, and binge drinking occasions and found that binge drinking is the most predictable phenotype of alcohol misuse. Similarly, we also compared different ML models and confound-control techniques and found that the two non-linear models - SVM-rbf and GB - perform better than the two linear models, SVM-lin and LR. Among the confound-control techniques, we found that counter-balancing with oversampling is most beneficial for the task. To the best of our knowledge, this was the first study to analyze and report results on the follow-up 3 data from IMAGEN. The results of our exploratory study advocate that collecting long-term, large cohorts of data, representative of the population, followed by a systematic ML analysis can greatly benefit research on complex psychiatric disorders such as AUD.

## Data Availability

All data produced are available online at https://github.com/RoshanRane/ML_for_IMAGEN

## AUTHOR CONTRIBUTIONS

Roshan Prakash Rane, Evert Ferdinand de Man, and Kerstin Ritter designed the study. Evert Ferdinand de Man preprocessed the data. Evert Ferdinand de Man and Roshan Prakash Rane performed the data analysis. JiHoon Kim contributed to the feature visualization. Roshan Prakash Rane, Kerstin Ritter, Evert Ferdinand de Man, Kai Görgen, Mira Tschorn, Henrik Walter, and Andreas Heinz prepared the manuscript. All other co-authors performed data acquisition and revised the manuscript.

## ACKNOWLEDGMENTS

We acknowledge support from the German Research Foundation (DFG, 389563835; 402170461-TRR 265; 414984028-CRC 1404; EXC 2002/1 “Science of Intelligence” – project number 390523135), the Brain & Behavior Research Foundation (NARSAD grant) and the Manfred and Ursula-Müller Stiftung. Gunter Schumann is a recipient of an Alexander von Humboldt Preis and a National Science Foundation of China (NSFC) Research Fund for International Scientists (Grant No. 82150710554).

## Footnotes

1 *https://imagen-europe.com/standard-operating-procedures*

2 *https://github.com/imagen2/imagenprocessing/tree/master/fsldti*

3 *AUDIT questionnaire (link)*

4 *https://github.com/RoshanRane/ML for IMAGEN*

5 *https://scikit-learn.org/stable/about.html*

