## Supplementary_material for "Structural differences in adolescent brains can predict alcohol misuse"

### 1 LITERATURE REVIEW

**Table S1. Overview of the literature** that looks at structural differences in the brain of adolescent alcohol misusers (AAMs) and healthy controls. The studies are sorted by the year of publication. For each study, the sample size 'n', the main analysis technique, and the main structural differences found in AAMs are listed.

| Study (year) | n | Analysis / method | Structural differences in AAMs |
|---|---|---|---|
| De Bellis et al. (2000) (2000) | 36 | Statistically compare (univariate) regional brain volumes between groups | Lower hippocampal volume. |
| Nagel et al. (2005) (2005) | 31 | Statistically compare (univariate) regional brain volumes between groups | Lower volume only in left hippocampus after controlling for other psychiatric comorbidities. |
| De Bellis et al. (2005) (2005) | 42 | Statistically compare (univariate) regional brain volumes between groups | Lower pFC, cerebellum volumes in males but AAMs had comorbid mental disorders. |
| McQueeny et al. (2009) (2009) | 28 | Mass-univariate analysis of skeletonized FA voxels (DTI) | Binge drinkers had lower FA in 18 white matter areas. |
| Squeglia et al. (2012) (2012) | 59 | Statistically compare (univariate) regional brain volumes between groups | No effect of binge drinking on cortical thickness and sex-specific differences among AAMs in left frontal cortex. |
| Jacobus et al. (2013) (2013) | 54 | Mass-univariate analysis of skeletonized FA voxels (DTI) | No effect in AAM-only group, but lower FA in AAM and comorbid marijuana users. |
| Luciana et al. (2013) (2013) | 55 | Longitudinal mass-univariate analysis of cortical thickness, white matter extent, DTI-extracted FA and MD | Accelerated GM thinning in mid frontal gyrus, attenuated WM growth with lower FA in left caudate, thalamus. |
| Whelan et al. (2014) (2014) | 692 | Exploratory analysis using ML to find best predictors of AAM among demographic, psychosocial, genetic, cortical volumes, and fMRI variables | Current AAMs have lower GMVs in parts of frontal lobe and higher GMV in right putamen. Future AAMs have lower GMV in right parahippocampal gyrus and higher in left postcentral gyrus. |
| Squeglia et al. (2017) (2014) | 137 | Exploratory analysis using ML to find best predictors of AAM among demographic, neuropsychological, cortical thickness, and fMRI variables | Future AAM have thinner GM in precuneus, lateral occipital, ACC, PCC, and frontal and temporal cortex. |
| Pfefferbaum et al. (2018) (2017) | 483 | Longitudinal mass-univariate analysis of GMV development | Accelerated GMV reduction in frontal brain regions. |
| Jones and Nagel (2019) (2019) | 113 | Modeling the WM microstructure development (DTI) for each voxel | Altered frontostriatal WM microstructure is predictive of future AAM. |
| Kühn et al. (2019) (2019) | ≈ 1500 | Growth curve modeling of GM volumes | Higher GMV in caudate nucleus and left cerebellum predicts future AAMs |
| Seo et al. (2019) (2019) | ≈ 1000 | ML analysis of cue-related brain region followed by mass-univariate analysis for identifying region importance | Current AAMs show reduced GMV in medial-pFC, oFC, thalamus, bilateral ACC, left amygdala and anterior insular. |
| Sullivan et al. (2020) (2020) | 548 | Longitudinal mass-univariate (GLM) analysis of cerebellar region volumes | Cerebellum: accelerated GM decline in in 2 sub-regions and accelerated expansion of WM in one sub-region and CSF. |
| Robert et al. (2020) (2020) | 726 | Mass-univariate analyses of voxels, followed by analysis of the direction of causality using causal bayesian networks | Accelerated GM atrophy in parts of the temporal cortex and left prefrontal cortex. |

Acronyms::: GM:grey matter; WM:white matter; CSF-cerebrospinal fluid; GMV:grey matter volume; pFC:prefrontal Cortex; oFC:orbitofrontal cortex; ACC:anterior cingulate cortex; PCC:posterior cingulate cortex; GLM:generalized linear models; ML:machine learning; DTI:Diffusion Tensor Imaging; FA:Fractional Anisotropy; MD:mean diffusivity

### 2 AAM PHENOTYPES

| Phenotype | IMAGEN questionnaire | Total range | Safe users *range (n)* | Moderate misusers *range (n)* | Heavy misusers *range (n)* |
|---|---|---|---|---|---|
| Frequency | ESPAD 8b | 0-6 | 0-4 (397) | 5 (270) | 6 (372) |
| Amount | AUDIT q2 | 0-4 | 0 (413) | 1 (403) | 2-4 (219) |
| Onset | ESPAD 29d | 11-21 | 16-21 (531) | 14-15 (288) | 11-14 (216) |
| Binge | ESPAD 19a | 0-6 | 0-3 (299) | 4-5 (336) | 6 (400) |
| Binge-growth | Growth curve of ESPAD 19b | 0-9 | 0-2 (379) | 3-5 (420) | 6-9 (236) |
| AUDIT | AUDIT-total | 0-40 | 0-4 (443) | 5-7 (274) | 8-40 (318) |
| AUDIT-quick | AUDIT-freq | 0-12 | 0-3 (402) | 4-5 (359) | 6-12 (274) |
| AUDIT-growth | Growth curve of AUDIT-total | 0-6 | 0,3 (377) | 4 (404) | 2,5,6 (254) |
| Combined-seo | ESPAD 8b, 17b, 19b, and TLFB alcohol2 | 0-2 | 0 (345) | 1 (404) | 2 (286) |
| Combined-ours | AUDIT q1, q2, ESPAD 19a, growth curve of ESPAD 19b | 0-3 | 0 (429) | 1 (403) | 2 (203) |

**Table S2.** The table explains how the ten AAM phenotypes are derived from the respective IMAGEN questionnaire. It lists the total values in that question and what range of values are used to categorize the subjects into safe users, moderate users and heavy users, respectively. For reference, the sample sizes ($n$) obtained at FU3 by using these value ranges are also shown in the brackets.

### 3 MACHINE LEARNING HYPERPARAMETERS

Each of the machine learning (ML) models have their own specific set of hyperparameters that are tuned using a 5-fold inner cross-validation during the exploratory analysis stage as shown in the table below. For both $C$ and $\gamma$, higher values lead to overfitting and lower values can lead to underfitting. For gradient boosting, the maximum depth of the trees is set at $5$, the maximum numbers of estimators at $100$, and the subsampling of input features is disabled as counterbalancing is used. The remaining parameters are set at the default values as defined in the *scikit-learn* python package[1].

[1] *https://scikit-learn.org/stable/about.html*

| Model | hyperparameter | values tested |
|---|---|---|
| Logistic regression | $C$: Inverse of $L2$ regularization strength | 1000, 100, 1.0, 0.001 |
| Linear support vector machine | $C$: Inverse of $L2$ regularization strength | 1000, 100, 1.0, 0.001 |
| Kernel-based support vector machine | $C$: Inverse of $L2$ regularization strength | 1000, 100, 1.0, 0.001 |
| | $\gamma$: kernel coefficient of RBF kernel | 'auto', 'scale' |
| Gradient boosting | learning_rate | 0.05, 0.25 |

### 4 ALGORITHM

**Algorithm 1** Procedure followed for each of the 3 analyses. The '$\models$' operation represents fitting or training the ML model given on the left side of the operation on the data given on the right side:

$\{data_{explore}, data_{holdout}\} \subset data_{infer}$ ▷ Keep aside 20% as $data_{holdout}$

**Start** exploratory analysis

$M \in \{$LR, SVM-lin, SVM-rbf, GB$\}$
$y \in \{y_{freq}, y_{amount}, ...y_{binge}\}$ ▷ select one of 10 AAM phenotypes

**for** $i_{outer} \in \{1, 2, ..., 7\}$ **do** ▷ Split $data_{explore}$ into 7 equal outer folds
    $train_{outer} \leftarrow \{data_{explore}[i] \mid i \neq i_{outer}\}$
    $test_{outer} \leftarrow \{data_{explore}[i] \mid i = i_{outer}\}$

    **for** $P \in \mathbb{P}$ **do** ▷ $\mathbb{P}$ is set of all hyperparameter combinations
        **for** $i_{inner} \in \{1, 2, ..., 5\}$ **do** ▷ Split $train_{outer}$ into 5 equal inner folds
            $train_{inner} \leftarrow \{train_{outer}[i] \mid i \neq i_{inner}\}$
            $test_{inner} \leftarrow \{train_{outer}[i] \mid i = i_{inner}\}$
            $M(P) \models train_{inner}$
            $acc_i =$ evaluate$(M(P), test_{inner})$
        **end for**
        $acc_P =$ mean$(acc_i \mid \forall i_{inner})$ ▷ average accuracy for hyperparameter combination $P$
    **end for**

    $\hat{P} \leftarrow \{P \mid$ highest$(acc_P \mid P \in \mathbb{P})\}$
    $M(\hat{P}) \models train_{outer}$
    $acc_j =$ evaluate$(M(\hat{P}), test_{outer})$
**end for**

$acc_{(M,y)} =$ mean$(acc_j \mid \forall i_{outer})$ ▷ average accuracy for model $M$ and label $y$
$\hat{M}, \hat{y} \leftarrow \{M \mid$ highest$(acc_{(M,y)} \mid \forall (M, y))\}$ ▷ select the best model $\hat{M}$ and AAM phenotype $\hat{y}$

**Start** generalization test

$\hat{M}(\hat{P}) \models data_{explore}$
$acc =$ evaluate$(\hat{M}(\hat{P}), data_{holdout})$

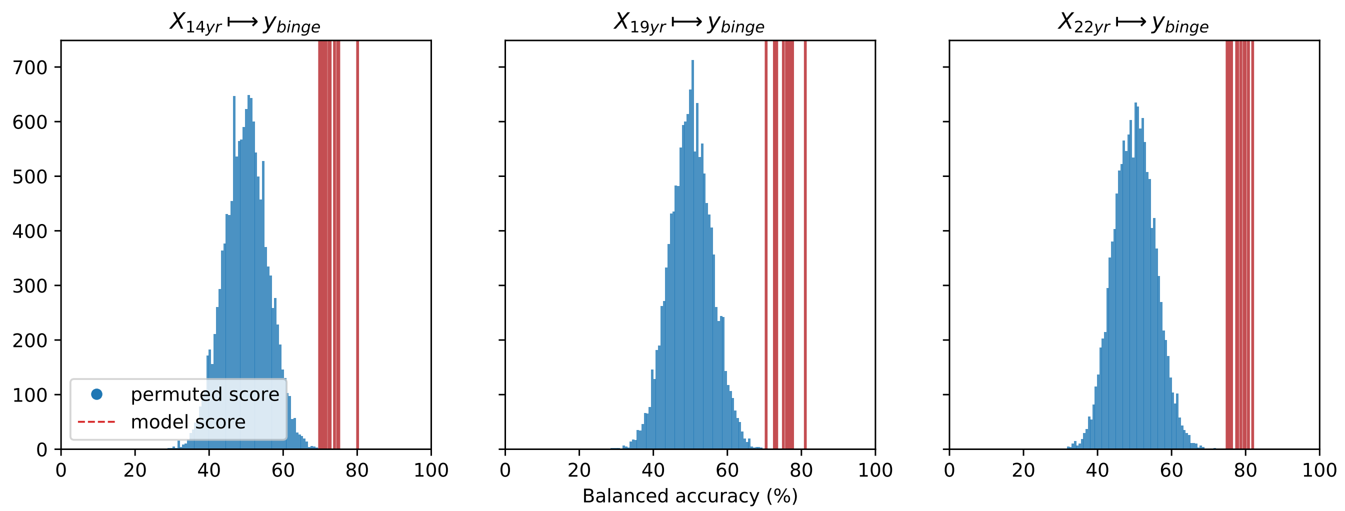


**Figure S1.** Distribution of model accuracy versus accuracy from 1000 random permutation tests

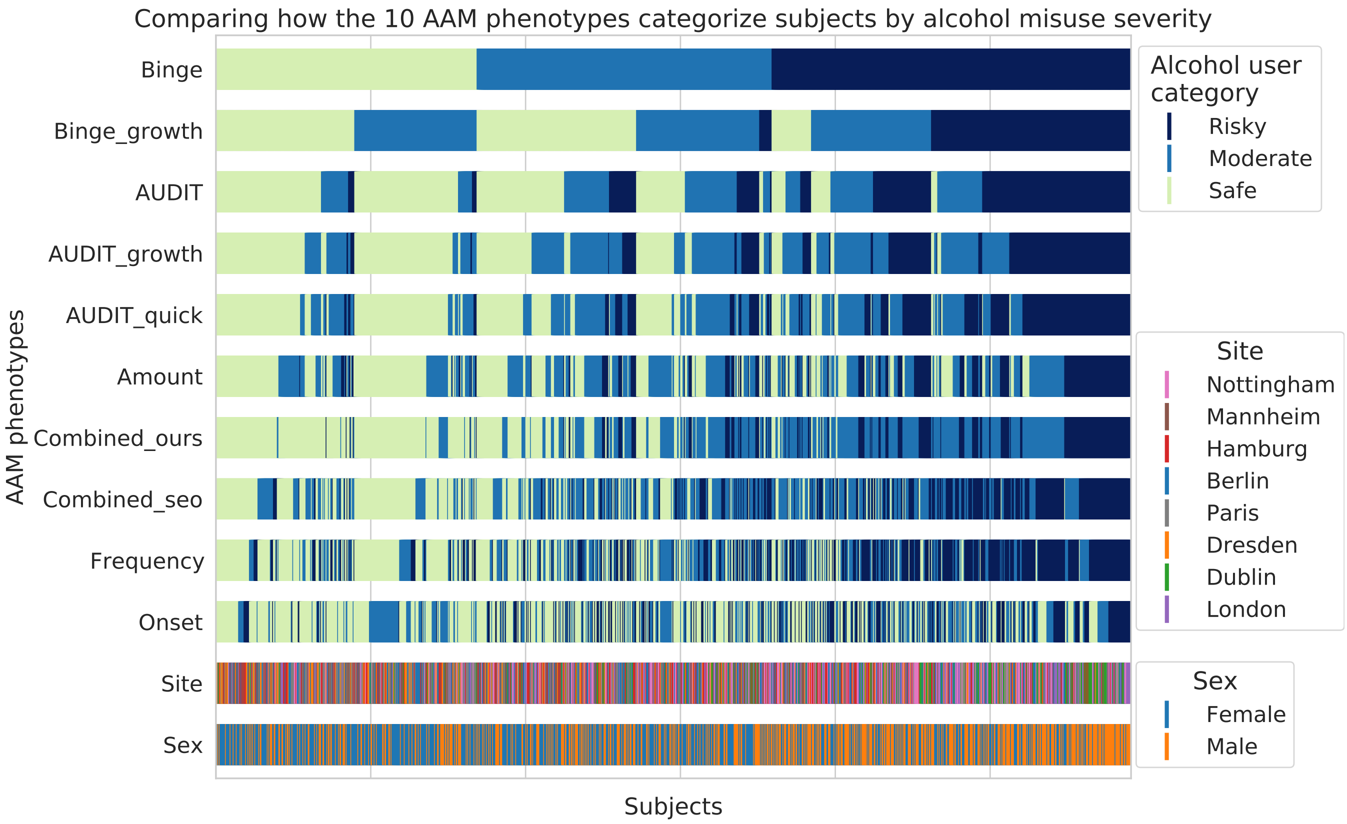


**Figure S2.** A qualitative comparison showing how the ten AAM phenotypes categorize the same subjects into the three alcohol user classes – risky alcohol users, moderate users, and safe or non-users. Each color-coded vertical line in the diagram represents one subject, out of the total 1182 subjects. It can be observed that the Frequency, Onset, and Amount phenotypes categorize very differently from Binge, showing that they capture different factors of alcohol misuse. All AUDIT-derived phenotypes are similar to each other but are different from the Binge phenotype. Furthermore, sex and site-based differences are detectable. Most males are categorized in the risky group compared to females and most subjects from Dublin are categorized as the 'risky' group by the Binge phenotype.

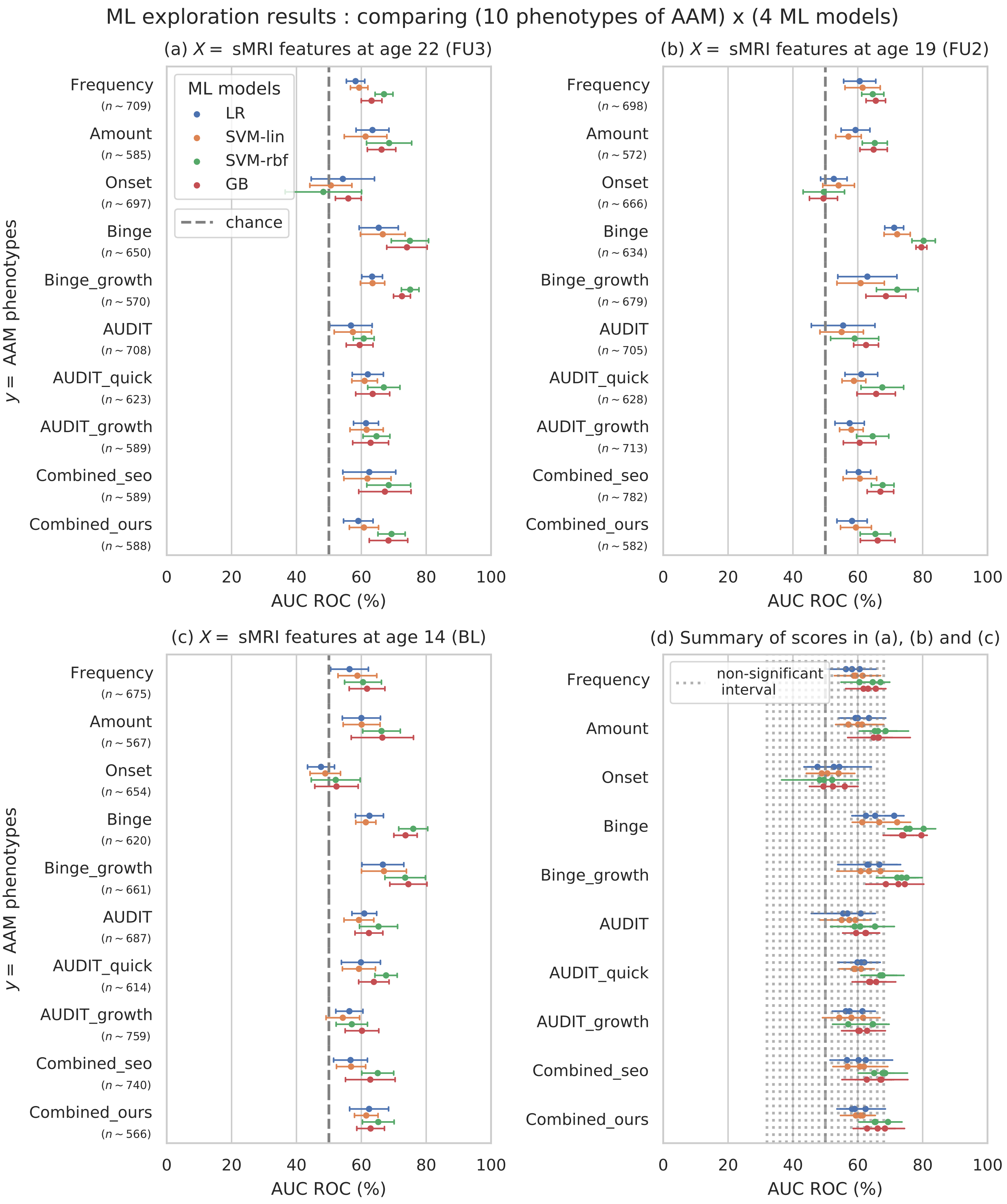


**Figure S3.** Results of the exploratory analysis with AUC-ROC metric

### 5 EXTENDED RESULTS

#### Cross-site experiment

Since multi-site data is used, an additional experiment is performed to test the ability of the ML models to generalize across recruitment sites. In this experiment, instead of randomly sampling $20\%$ of the subjects as the data$_{holdout}$, all the subjects from the Nottingham site are set aside that has approximately $20\%$ of all subjects ($n = 176$). For the 7-fold CV performed in the ML exploration, subjects from each of the remaining 7 recruitment site are used as one fold. This method of CV is termed *leave-one-site-out* CV Rozycki et al. (2018). The result from the *leave-one-site-out* CV experiment is reported in Figure S4. The ML models fail to produce a significant performance for any of the three time points in the final generalization test and perform close to chance for all AAM labels in the ML exploration experiments.

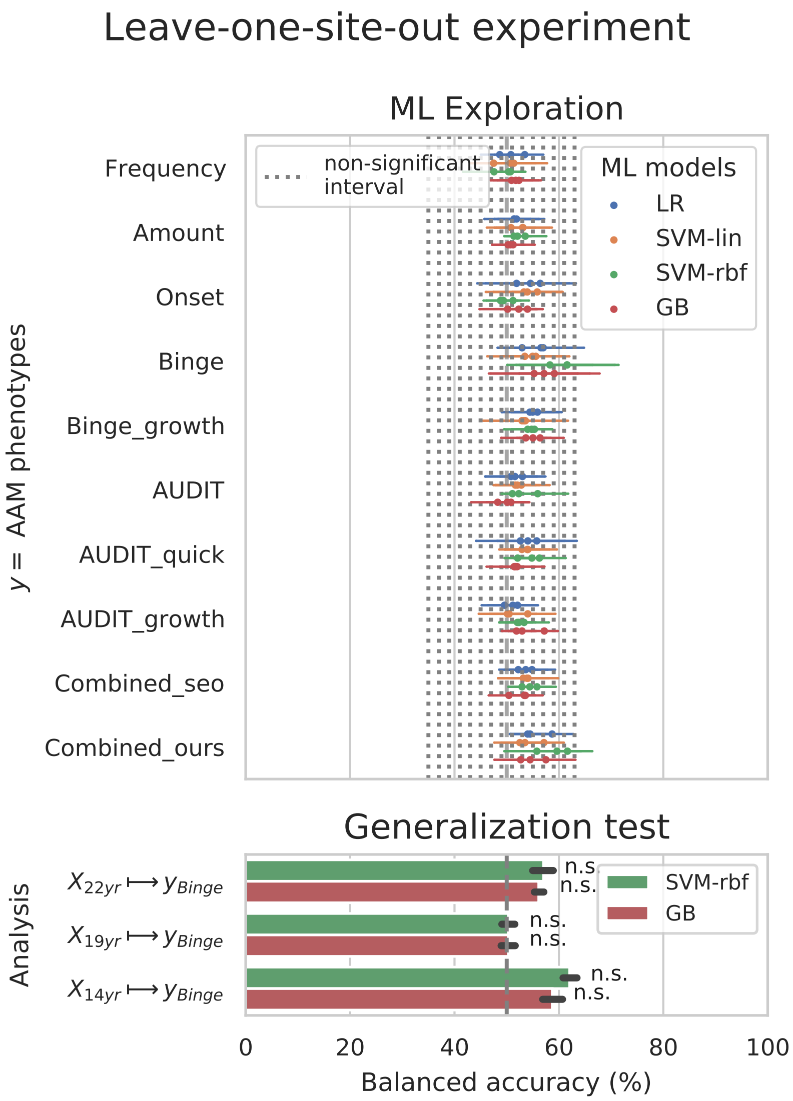


**Figure S4.** Analysis repeated with leave-one-site-out cross validations (CV).
